# Association between Insomnia Symptoms and Risk of Heart Failure: A Meta-analysis of Prospective Cohort Studies

**DOI:** 10.1101/2024.03.20.24304651

**Authors:** Juan Gao, Le Han, Hong-Shuai Cao, Xiao-Qin Luo, Li-Yue Xu, Ying Zhou, Zhe-Xun Lian, Jing-Yi Ren

**Affiliations:** Department of Cardiology, The Affiliated Hospital of Qingdao University, Qingdao, China; Heart Failure Center, Department of Cardiology, China-Japan Friendship Hospital, Beijing, China; Department of Respiratory Medicine, China-Japan Friendship Hospital, Beijing, China

**Keywords:** Insomnia symptoms, DIS, Heart failure events, Meta-analysis, Prospective cohort studies

## Abstract

**Background:** Insomnia, as one of the most prevalent sleep disorders, is frequently linked to Heart failure (HF). However, the precise relationship and potential risk of HF events associated with insomnia and subtypes of symptoms necessitate further investigation.

**Objective:** This updated meta-analysis aimed to evaluate the associations between HF and various insomnia symptoms, including difficulty initiating sleep (DIS), difficulty maintaining sleep (DMS), early-morning awakening (EMA) and non-restorative sleep (NRS).

**Methods:** A comprehensive literature search was conducted in PubMed, Web of Science, Embase, and the Cochrane Library to identify prospective cohort studies from inception to October 2023. Pooled hazard ratios (HRs) with 95% confidence intervals (CIs) were calculated to assess the correlation between insomnia and the risk of HF. Funnel plots and Egger’s tests were employed to assess publication bias.

**Results:** A total of 177,008 patients from seven eligible prospective cohort studies were included. The pooled minimally adjusted HR for HF was 1.26 (95% CI:1.09-1.45; *P*=0.001), indicating that insomnia was associated with increased risk of HF. Among the individual insomnia symptoms, only DIS showed a positive association with risk of HF (HR:1.14, 95% CI:1.04-1.25; *P*=0.005). DMS and NRS had no significant effect on the risk of HF (*P*>0.05). In the subgroup analysis including age, sex and BMI, there were no significant differences between each group.

**Conclusion:** This meta-analysis confirms the link between insomnia and an increased risk of HF, particularly highlighting the importance of DIS as a potential predictor for HF.

**Clinical Perspective:** What is new?

Insomnia is ubiquitous and seriously affects people’s normal work, life, and health. At present, prospective cohort studies have suggested that insomnia can increase the risk of heart failure (HF). However, in our meta-analysis, we firstly found that only Difficulty Initiating Sleep (DIS) was directly associated with an increased HF incidence, rather than other insomnia subtypes. This is a very interesting which suggests that DIS has a greater impact on the incident of HF. In addition, this points us that in the subsequent treatment of insomnia, maybe we should pay more attention to the treatment of DIS.

What are the clinical implications?

As insomnia symptoms can be assessed and identified early and are relatively manageable, the study of the relationship between insomnia and HF has potential important clinical significance. If further research confirms that the improvement of insomnia symptoms, especially DIS symptoms, contributes to the prevention of HF or the prognosis of HF patients, it will provide another new “track” for the prevention and treatment of HF.

## Introduction

Sleep disturbances have become increasingly prevalent in modern society, with substantial implications for public health ^1^. Previous studies have confirmed that sleep apnea, including obstructive sleep apnea (OSA) and central sleep apnea (CSA), is independently associated with a higher risk of heart failure (HF) related symptom progression, hospitalization, and mortality. Insomnia, one of the most common sleep disorders, affecting ~10–15% of the population and having long-term adverse cardiovascular consequences ^2–4^. However, it is largely unknown whether insomnia is prospectively associated with an increased incidence of HF. In the context of HF, insomnia has gained attention as a significant comorbidity, with studies reporting a high prevalence of insomnia symptoms among HF patients ^5,6^. Common symptoms of insomnia including difficulty initiating sleep (DIS), difficulty maintaining sleep (DMS), early-morning awakening (EMA) and non-restorative sleep (NRS) have been reported in a substantial proportion of individuals with HF, ranging from 23% to 73% ^7–9^. A recent study indicates that these different subtypes may have different impact on cardiovascular disease ^10,11^. The prevalence of insomnia subtypes is related to demographic characteristics, anxiety, depression, alcohol consumption, and use of hypnotics ^12,13^(12,13). However, the precise nature of the association between different subtypes of insomnia and the subsequent risk of developing HF among individuals without HF at baseline remains poorly understood.

To address this knowledge gap, we conducted a comprehensive systematic review and meta-analysis to evaluate the potential impacts of insomnia and its different subtypes on the risk of incident HF. By systematically synthesizing the available evidence from prospective cohort studies, we aimed to provide insights into the role of insomnia as a potential risk factor for the incidence of HF. Our findings may have implications for early identification, intervention, and management strategies targeting insomnia to mitigate the risk of HF and improve cardiovascular outcomes.

## 1. Methods

### 2.1 Search strategy and selection criteria

This systematic review and meta-analysis were performed in accordance with the Cochrane Collaboration and Preferred Reporting Items for Systematic Reviews and Meta-Analyses (PRISMA) guidelines. We performed a systematic literature search in PubMed, Web of Science and EMBASE evaluating studies published up to October 9, 2023. The search string was: (‘heart’ AND ‘failure’) OR (‘heart’ AND ‘congestive’) OR (‘ventricular’ AND ‘dysfunction’) AND (‘insomnia’ OR ‘sleep disorders’ OR ‘sleep initiation and maintenance disorders’) AND (‘clinical trial’ OR ‘randomized’). And reference lists of review articles were also screened to ensure the inclusion of relevant studies. Search results were screened independently by two investigators (G.J. and H.L.) for the relevance of titles/abstracts and full texts of the studies fulfilling the inclusion criteria. Potential disagreements were solved by a reviewer (R.J). The systematic review was previously registered in PROSPERO (International Prospective Register of Systematic Reviews; Study Unique Identifier: 359352).

Studies were considered eligible if they fulfilled all the following criteria: (i) only studies with a prospective design were included to establish temporal relationships between insomnia symptoms and the risk of HF events; (ii) studies involving individuals aged 18 years or older were considered eligible for inclusion; (iii) reporting on the association between insomnia symptoms and the risk of HF events: Studies needed to report on the relationship between insomnia symptoms, such as DIS, DMS, EMA or NRS, and the occurrence of HF events; (iv) provision of hazard ratios (HRs) or sufficient data for their calculation: Studies were required to provide HRs, or data that allowed the calculation of HRs, along with corresponding 95% confidence intervals (CIs) or standard errors. Non-comparative studies, reviews, case-control studies, conference abstracts, meeting abstracts, comments, in vitro studies, animal studies, duplicate or continued work of previous publications and studies with incomplete data were excluded from the analysis.

### 2.2 Data extraction and quality assessment

Two investigators (G.J. and H.L.) independently extracted the following information from each study: name of the first author, publication year, geographical region, sample sizes, duration of follow-up, participant characteristics, definition and assessment of exposure and outcome, adjusted effect size (HRs with 95% CIs), and the covariates adjusted in the multivariable analysis. The Newcastle-Ottawa Scale (NOS) was used to assess the risk of bias in observational studies. The procedure was conducted by two independent investigators and disagreement was resolved by consensus. H.L. and L.X.Q performed risk of bias assessments. A cut-off of less than 5 points was considered as high risk of bias.

### 2.3 Statistical analysis

The heterogeneity between studies was assessed by the inconsistency index I^^2^ statistic (ranging from 0% to 100%) on the basis of the Cochrane Q test. Heterogeneity is considered to be low between studies if I^^2^ ranges from 0% to 25%, moderate from 25% to 75% and high from 75% to 100%. Pooled estimates HRs with 95% confidence interval was generated using Mantel-Haenszel transformation under the random effects models to evaluate the potential associations between insomnia symptoms and incidence of HF. A two-tailed *P*<0.05 was considered statistically significant. All statistical analysis were conducted using Review Manager software (version 5.3.5, Nordic Cochrane Centre, Copenhagen, Denmark).

## Results

### Literature search

The flowchart of the meta-analysis (Figure 1) shows the results of the published reports research. Of the 8,212 articles identified, we excluded 2,826 duplicate records, 4,806 articles were excluded due to nonrelevant articles (i.e., reviews, editorials, opinion letters, studies not written in English and original articles not matching the inclusion and exclusion criteria) or duplicate patient populations by screening of the title and abstract. 220 studies were excluded due to cell or animal studies. A total of 360 articles were evaluated as full-text articles, 353 articles were excluded for insufficient data or other types of diseases; Finally, 7 prospective cohort studies were deemed eligible for quantitative analysis.

**Figure 1.**
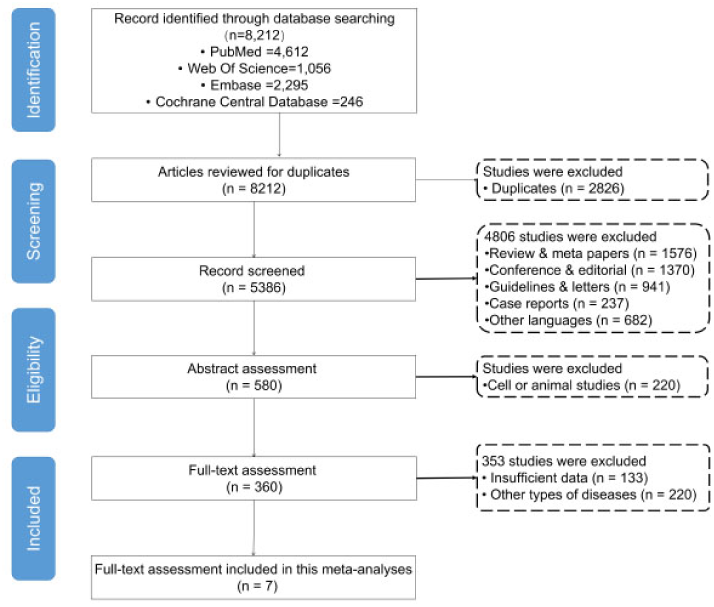
PRISMA flow diagram illustrating the screening and selection process: Schematic overview of study selection

### Study Characteristic

Although the present meta-analysis included 7 prospective cohort studies, the sample size of all participants was as high as 177,008, with the largest study including 54,279 participants and the smallest 2,314. This meta-analysis included representative countries from Europe, North America, and Asia. The age range of the participants varied from 20 to 79 years, and the follow-up duration ranged from 9 to 29.6 years. Meanwhile statistical analyses in each study involved the adjustment of several key variables to account for potential confounding factors. Especially, these studies describe the subtype symptoms of insomnia in patients, making it easier for us to conduct analysis of subtypes of insomnia. The main clinical characteristics of the included studies are summarized in Table 1. It provides comprehensive information on the first author, country of origin, study years, sample size, percentage of male participants, average age, average follow-up duration, and the number of HF cases reported in each study.

**Table 1.**
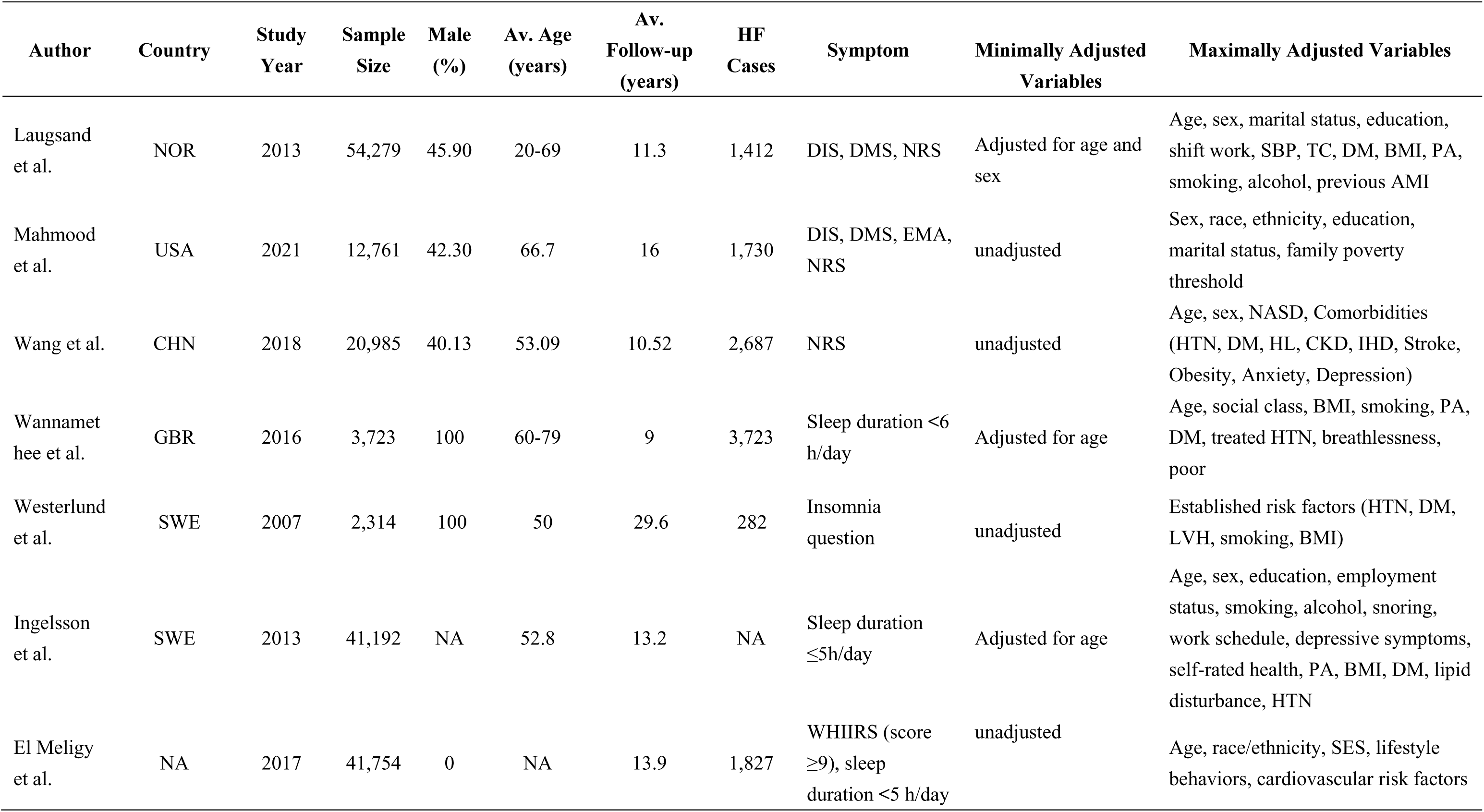
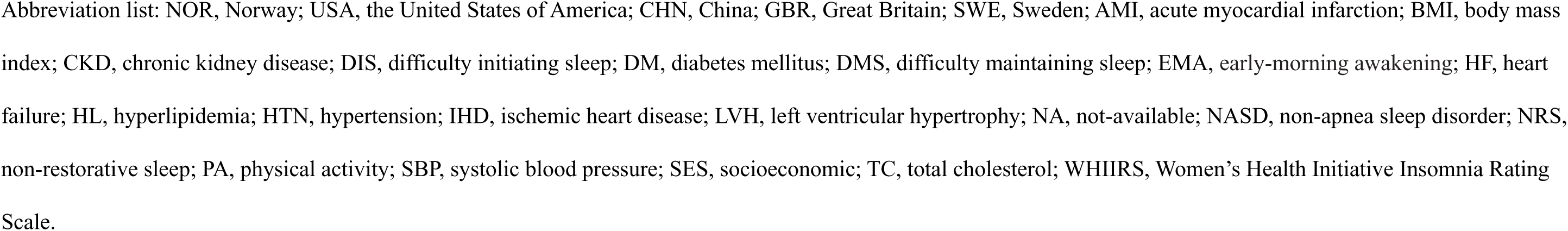
Summary of clinical characteristics in included studies.

### Meta-analysis results

#### Insomnia increases the risk of incident HF

The meta-analysis revealed significant insights into the association between insomnia and the risk of incident HF. Figure 2 illustrates the combined analysis results, demonstrating a substantial and statistically significant association between insomnia and incident of HF compared to the control group. The pooled minimally adjusted HR for HF, based on the combined analysis, was found to be 1.26 (95%CI:1.09-1.45, *P*=0.001; Figure 2A). This indicates that individuals with insomnia have a 26% higher risk of experiencing incident HF compared to those without insomnia. Furthermore, the maximally adjusted HR for HF was calculated to be 1.20 (95%CI:1.02-1.41, *P*=0.03; Figure 2B), providing additional support for the association between insomnia and risk of incident HF. The findings demonstrated a significant association between insomnia and risk of incident HF, indicating that insomnia may serve as a predictive factor for HF.

**Figure 2.**
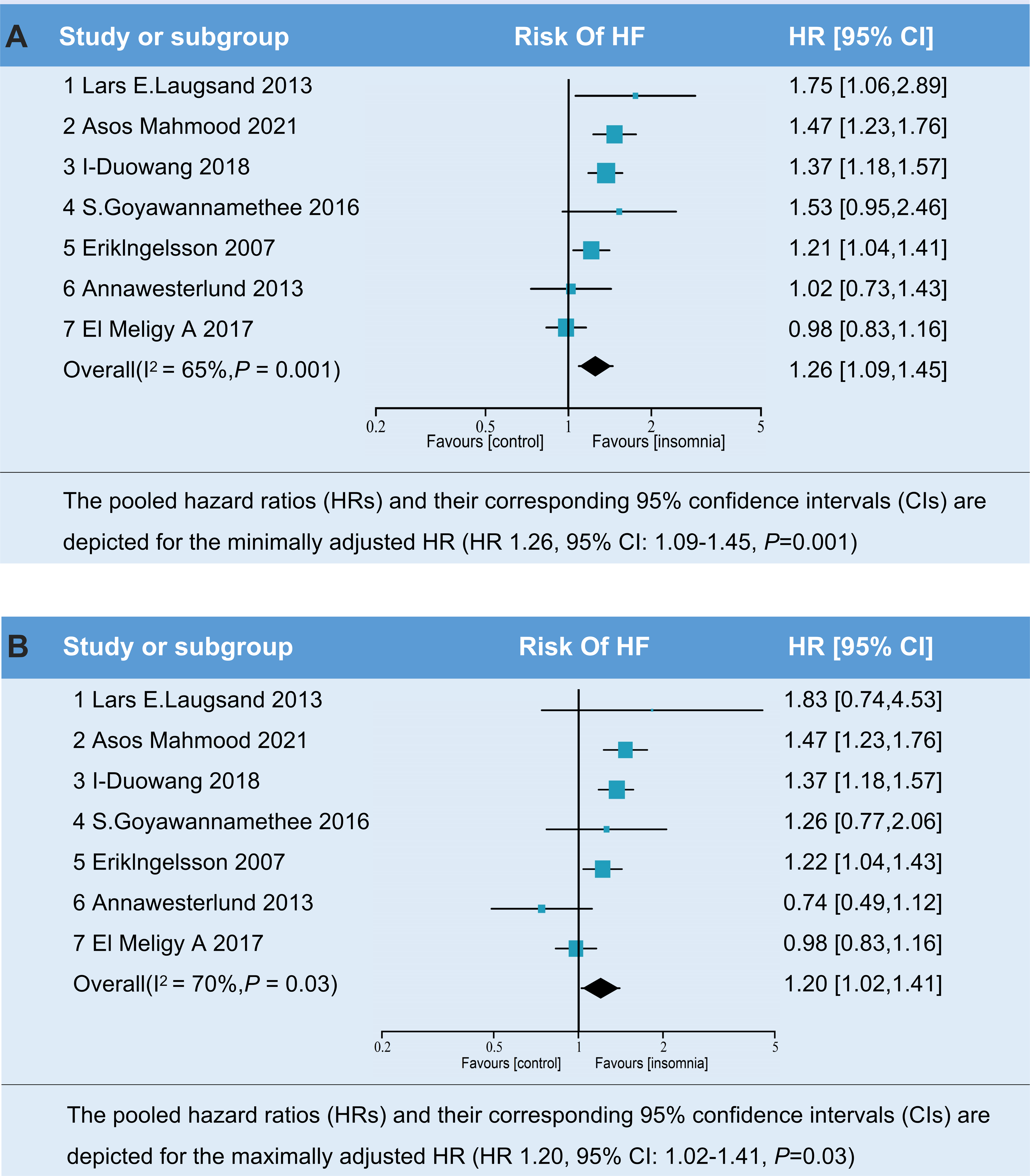
Forest plot of associations between insomnia and risk of HF: (A) The pooled hazard ratios (HRs) and their corresponding 95% confidence intervals (CIs) are depicted for the minimally adjusted HR (HR 1.26, 95% CI: 1.09-1.45, *P*=0.001). (B) The pooled hazard ratios (HRs) and their corresponding 95% confidence intervals (CIs) are depicted for the maximally adjusted HR (HR 1.20, 95% CI: 1.02-1.41, *P*=0.03).

#### DIS is the most important subtype of insomnia increased the risk of incident HF

Based on different clinical characteristics of various subtypes of insomnia ^14^(14), we analyzed the relationship between different subtypes of insomnia and the risk of incident HF, including DIS, DMS and NRS. Among the insomnia symptoms, the meta-analysis found that only DIS exhibited a statistically significant association with the risk of HF, with an HR of 1.14 (95% CIs:1.04-1.25, *P*=0.005; Figure 3A). For DMS and NRS, although the HR indicated a slight increase in the risk of HF, the associations were not statistically significant (HR:1.02, 95% CI:0.88-1.19, *P*=0.76; HR:1.02, 95% CI: 0.66-1.58, *P*=0.92, Figure 3B and 3C). Unfortunately, as only one study involved information of EMA, this meta-analysis was not included analysis of EMA and HF.

**Figure 3.**
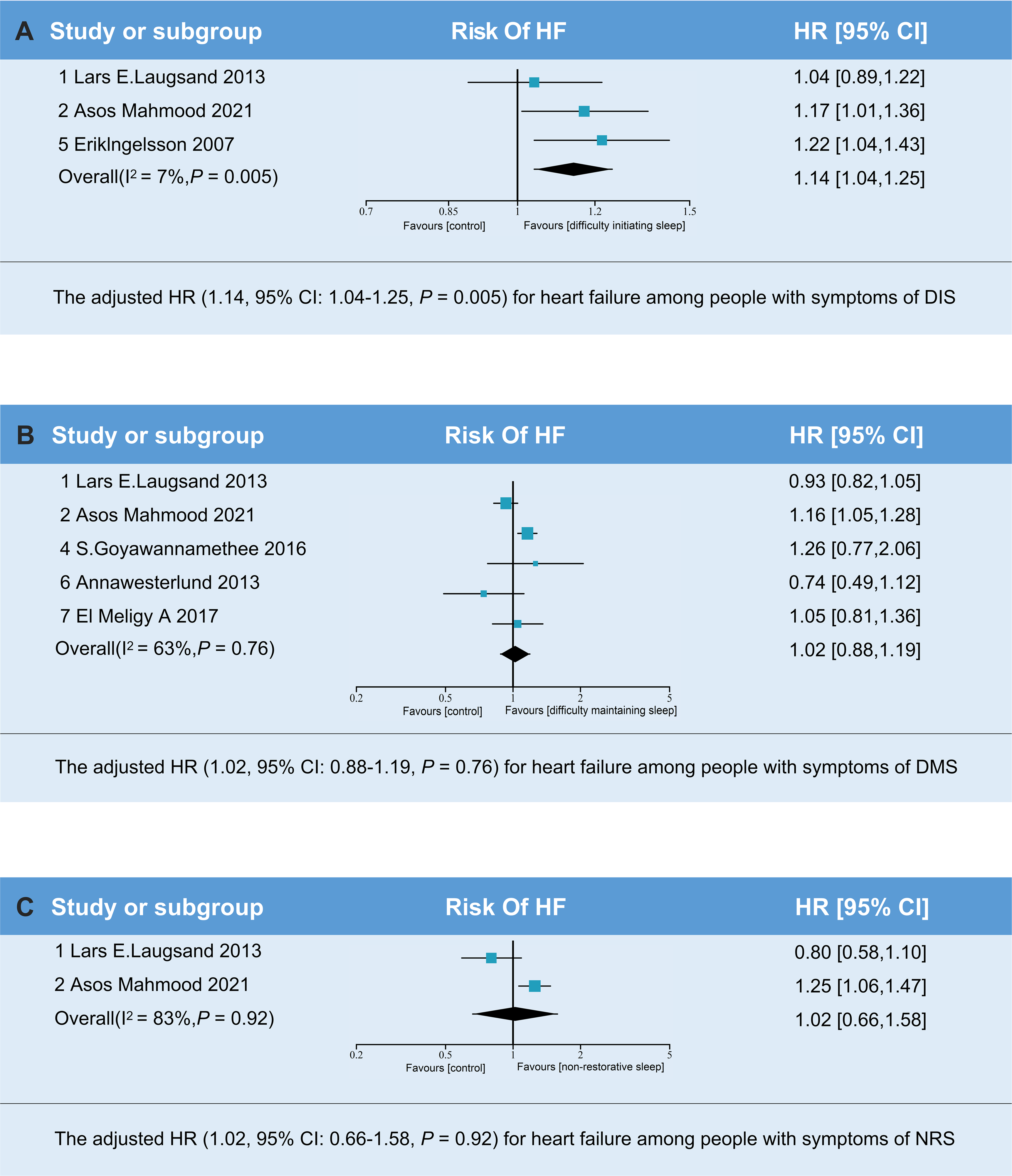
Forest plot of associations between subtypes of insomnia and risk of HF: (A) The adjusted HR (1.14, 95%CI:1.04-1.25) for HF among people with symptoms of DIS. (B) The adjusted HR (1.02, 95%CI:0.88-1.19) for HF among people with symptoms of DMS. (C) The adjusted HR (1.02, 95%CI:0.66-1.58) for HF among people with symptoms of NRS.

#### Subgroup analysis based on age, gender and BMI shows no difference

As is well known, elderly people and women are more prone to insomnia ^15,16^. Meanwhile, it is also common for overweight individuals to suffer from CSA ^17,18^. Thus, we assessed the impacts of these covariates on insomnia symptoms and risk of incident HF. Interestingly, we were surprised to found that there were no statistically significant differences between subgroups of age, gender and BMI (Figure 4). Further research is warranted to confirm the relationship and deeper pathogenesis between different covariates (such as age, gender and BMI) of insomnia and the risk of incident HF.

**Figure 4.**
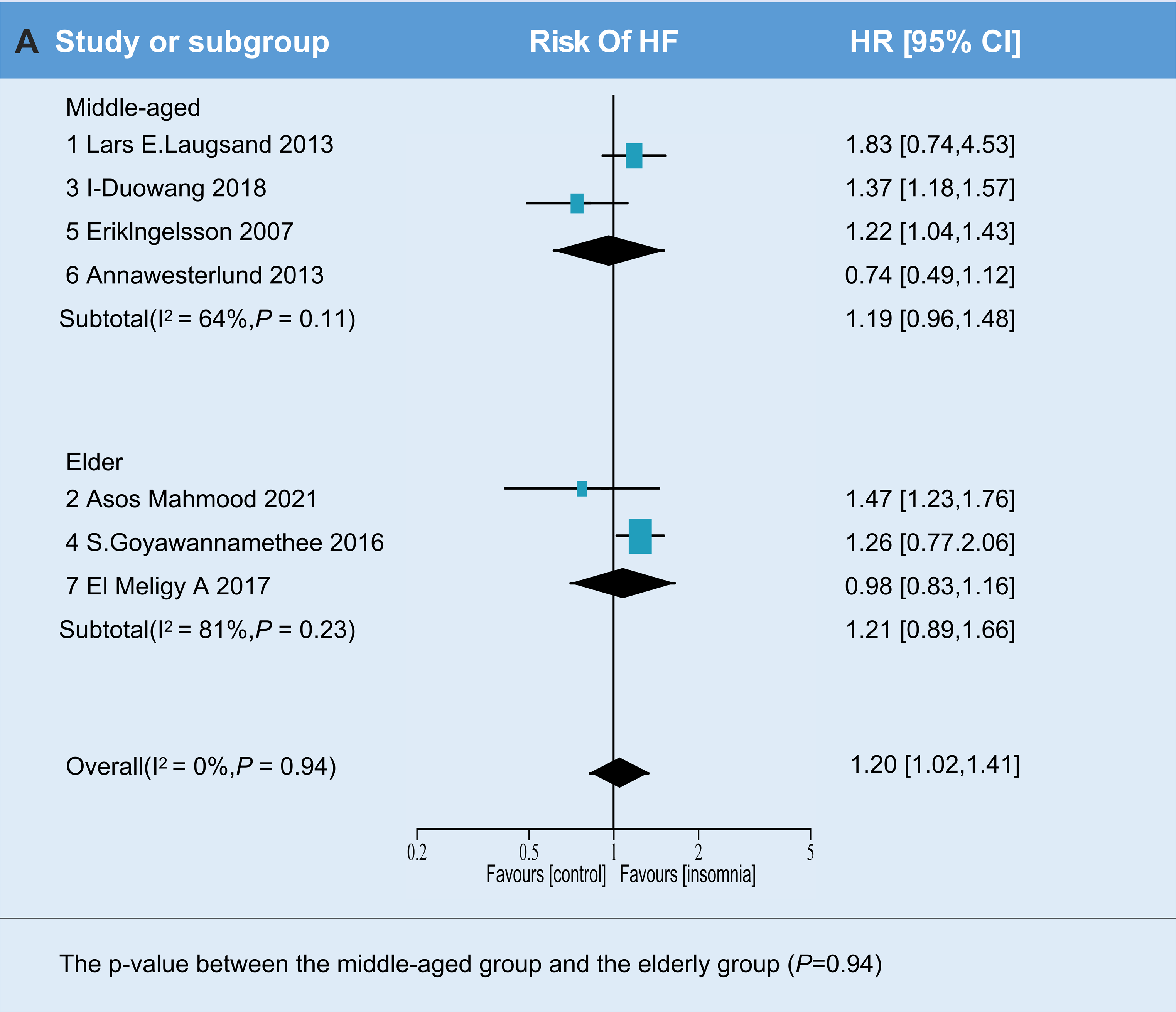
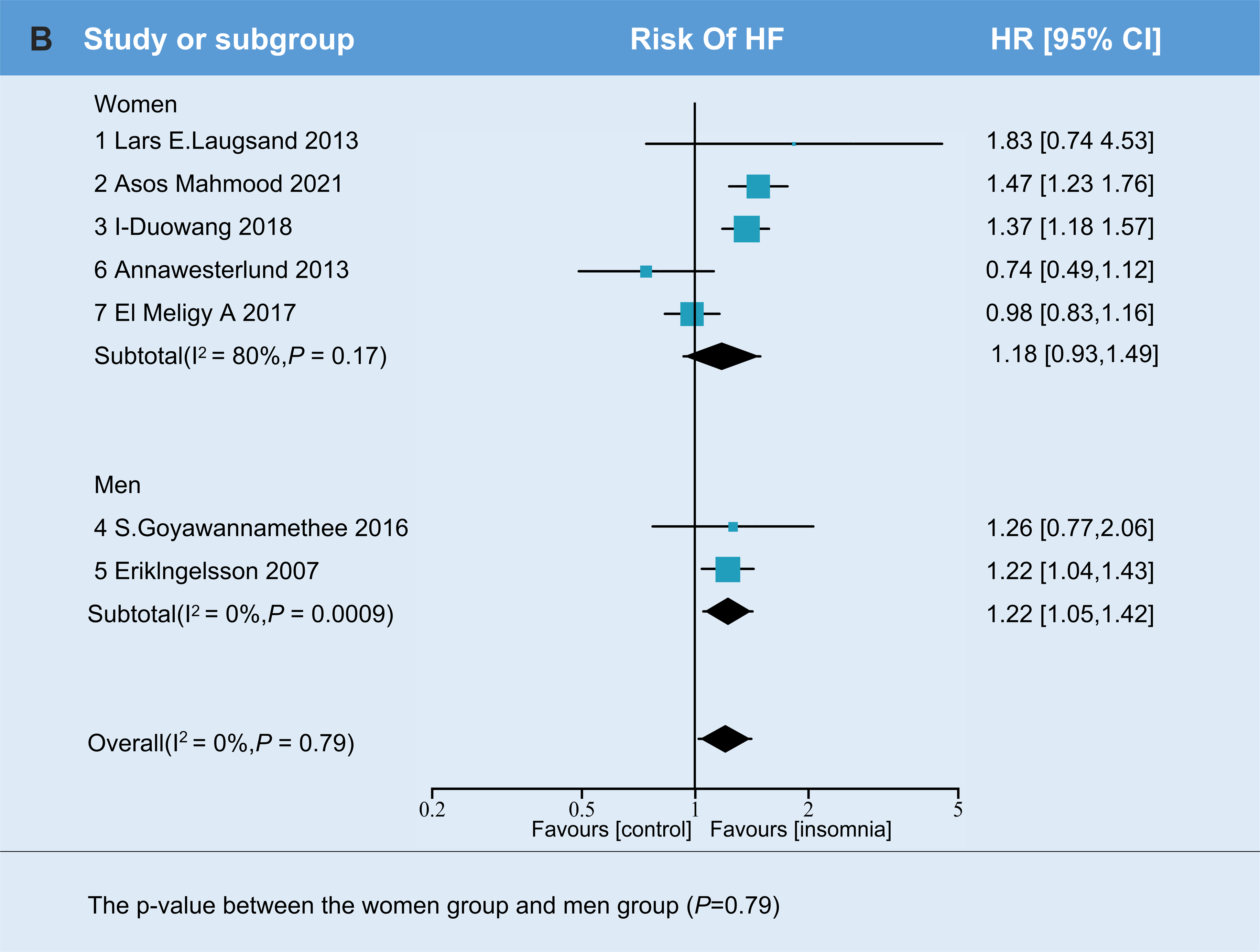
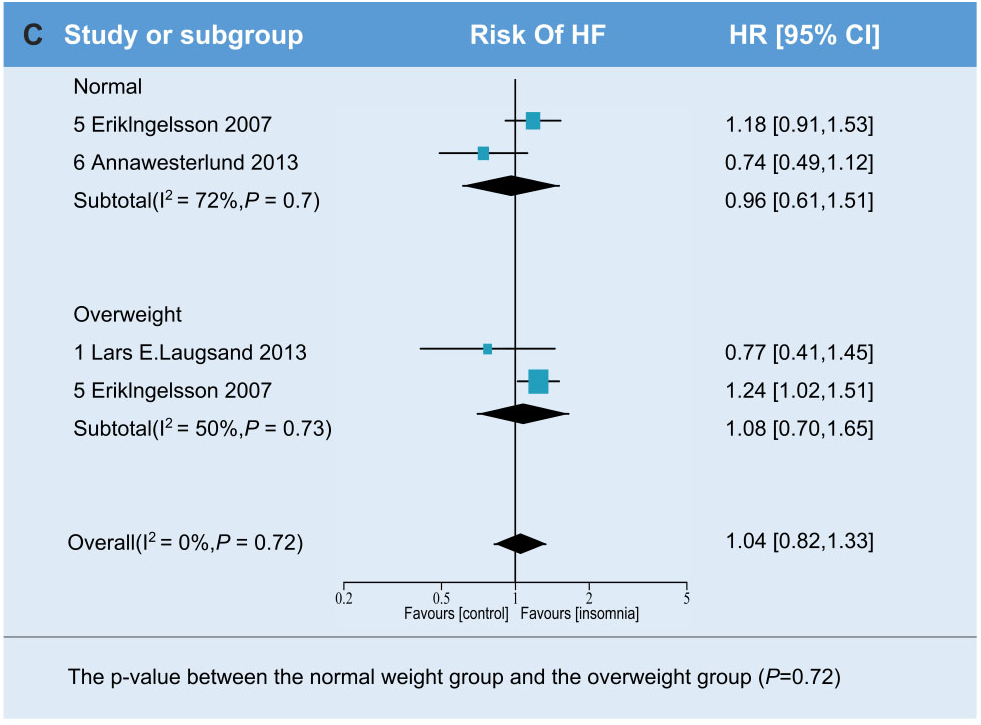
Subgroup analyses based on age, gender and BMI: (A) The p-value between the middle-aged group and the elderly group (*P*=0.94). (B) The p-value between the middle-aged group and the elderly group (*P*=0.79). (C) The p-value between the normal weight group and the overweight group (*P*=0.94).

#### Study quality and risk of bias assessment

Table 2 showed that all included studies with NOS scores (seven studies) receive 8 or more scores (mean=8.14), indicating relatively high methodological quality. Funnel plot analysis (Figure 5) was used to assess the potential publication bias in the included literature. The funnel plot shape, along with the results of Egger’s test, provided no statistical evidence supporting the presence of publication bias (*P*=0.868). The result showed that there were no notable indications of significant publication bias among studies.

**Figure 5.**
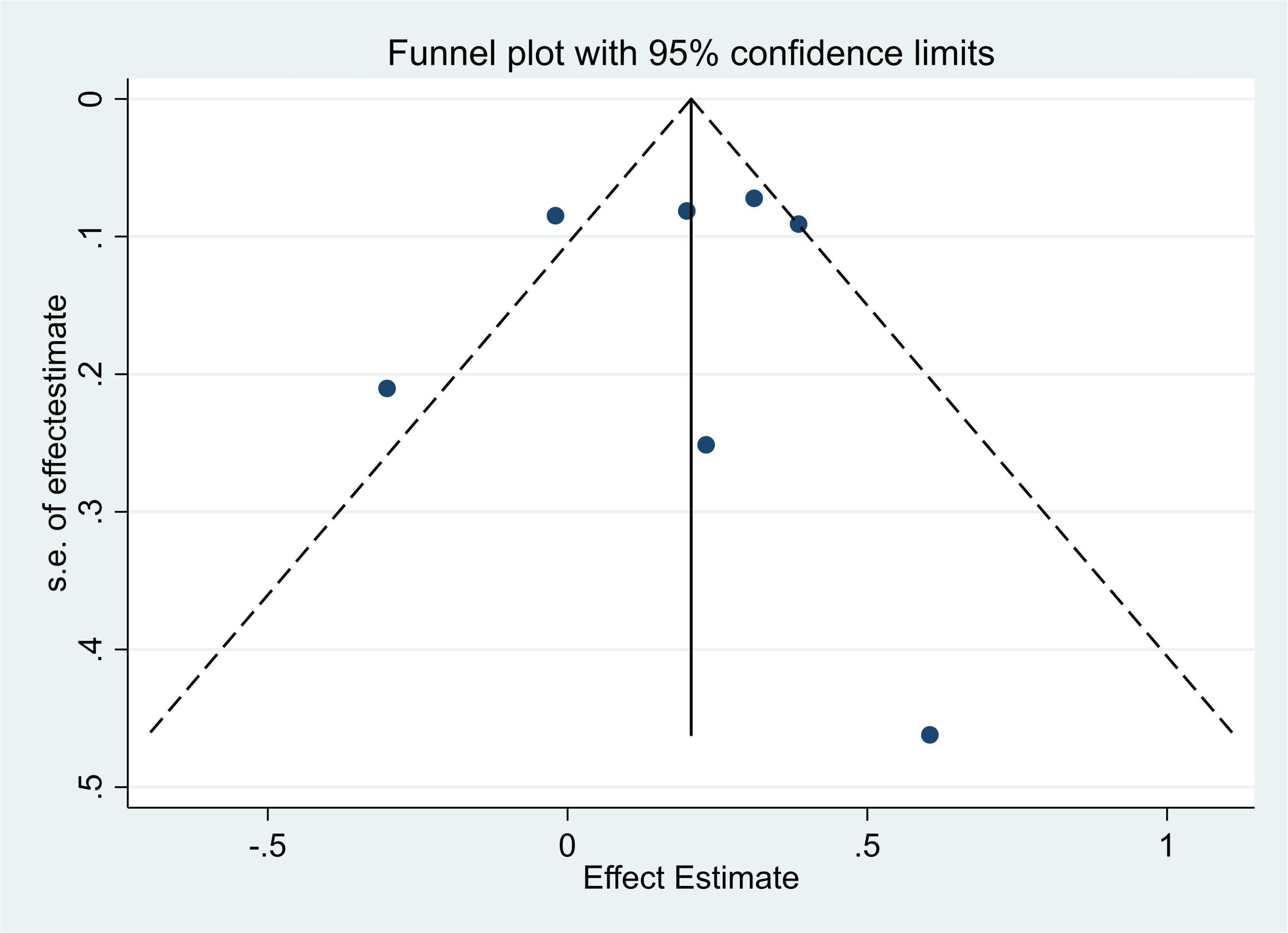
Funnel plot and Begg’s test for publication bias assessment: Egger test=0.868>0.05

**Table 2.**
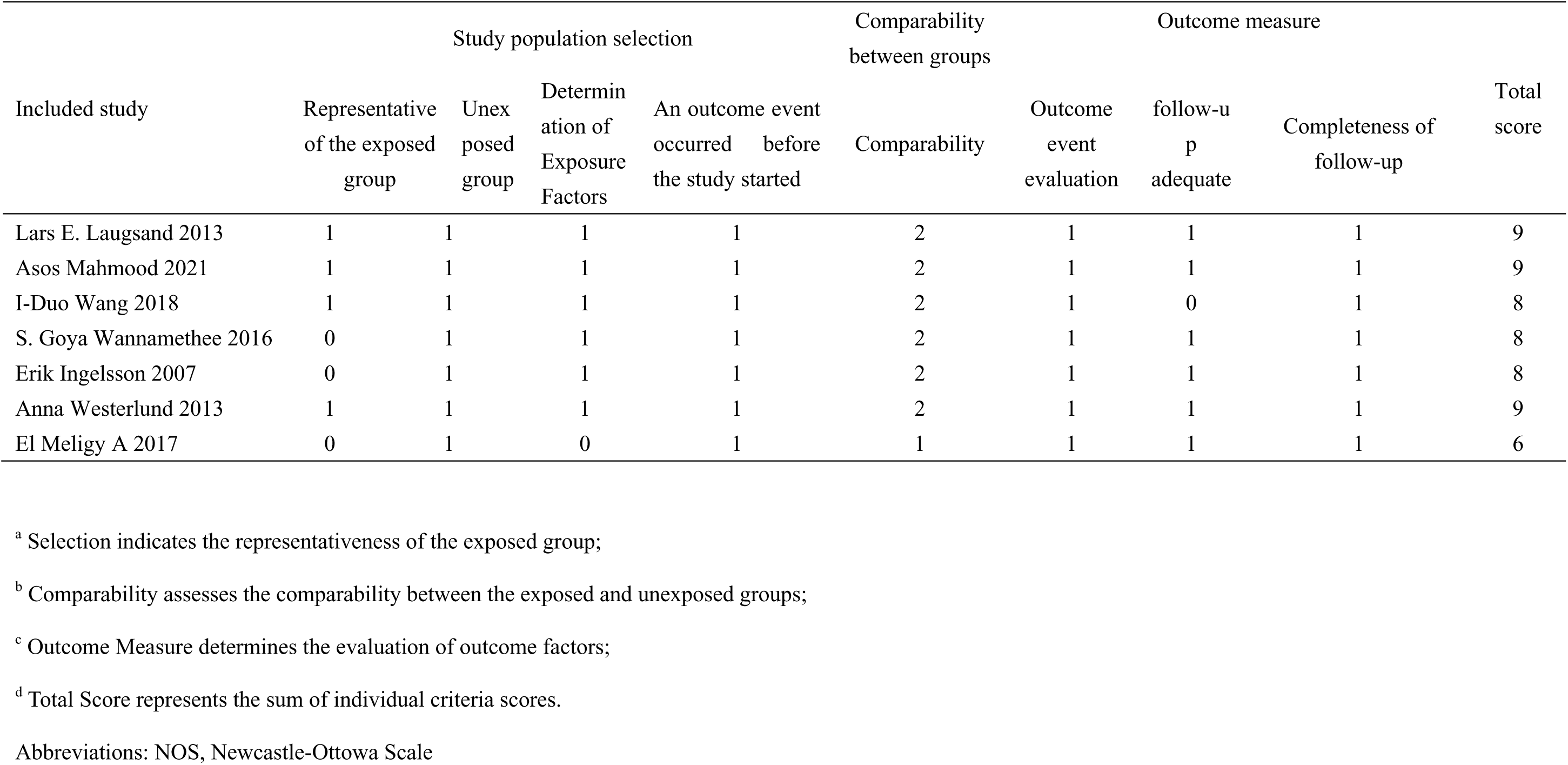
Newcastle-Ottawa Scale (NOS) assessment of included studies.

## Discussion

In this meta-analysis, we examined the association between insomnia and the risk of incident HF. We included 7 prospective cohort studies to investigate this relationship. This meta-analysis is the first, to our knowledge, to reveal the prospective association between insomnia and the risk of incident HF (HR=1.26, 95% CI: 1.09, 1.45; *P*=0.001). Based on the main characteristics of insomnia, we specifically looked at different subtypes of insomnia and interestingly found that only DIS, not DMS or NRS, was positively associated with the risk of developing HF. This suggests that DIS has a more significant impact on the risk of HF, which need more attention for clinicians.

The pathophysiology underlying insomnia and its association with HF has not yet fully understood. However, several mechanisms have been proposed to explain the link between insomnia-related symptoms and the incident of HF ^19,20^. The pathophysiology of insomnia and its impact on HF involves a complex interplay of neuroendocrine dysregulation, autonomic dysfunction, inflammation, and unhealthy behaviors ^21–23^. Firstly, in terms of the neuroendocrine system, insomnia is characterized by hyperarousal and chronic activation of stress responses, including increased activity in the hypothalamic-pituitary-adrenal (HPA) axis and the sympathetic nervous system ^24^. These results in elevated cortisol levels, upregulation of the renin-angiotensin-aldosterone system, and increased heart rate, decreased heart rate variability, and elevated blood pressure, all of which are known risk factors for HF ^25,26^. Secondly, in terms of inflammation, insomnia is associated with the increased secretion of pro-inflammatory cytokines and catecholamines, further contributing to the HF risk ^27^. Insomnia-related hypercortisolemia and sympathetic activation have also been implicated in insulin resistance and metabolic syndrome linked to HF pathophysiology ^28^. Furthermore, insomnia symptoms are associated with increased levels of pro-inflammatory biomarkers, which play an important role in HF development ^29^. Thirdly, maladaptive or inadequate health behaviors among individuals with insomnia, such as poor diet and physical inactivity, may also contribute to the incident of HF ^30^. Thus, dysregulation of the HPA axis, abnormal modulation of the autonomic nervous system, increased sympathetic activity, systemic inflammation, and unhealthy behaviors could potentially explain the association between insomnia symptoms and the incident of HF.

Among different subtypes of insomnia, DIS is characterized by delayed falling sleep, DMS is characterized by interruption of sleep continuity, while NRS is mainly characterized by poor sleep quality ^31^. Based on different subtypes of insomnia may lead to various cardiovascular outcomes, we specifically focused on subtypes of insomnia symptoms. Our meta-analysis interestingly found that only DIS, not DMS or NRS, was positively associated with the risk of HF. Although the underlying pathological mechanism remains unknown, it may be related to DIS increasing oxidative stress and inflammation in endothelial cells. One study showed that delaying bedtime by 1.5 h impairs clearance of endothelial oxidative stress that over time increases cardiovascular risk ^32^. The study identified reduced expression of DCUN1D3, a protein that facilitates Nrf2-mediated antioxidant response in ECs, as a novel mechanism mediating the lack of endothelial antioxidant response to sleep restriction induced oxidative stress ^33^. Thus, previous study has enlightening implications for us, for the main characteristic of DIS is difficulty falling asleep. This suggests that DIS has a greater impact on the incident of HF, which highlights the need of additional attention of clinicians. However, its deeper mechanisms require further exploration.

As is well known, elderly people are more prone to insomnia and HF ^15^. However, when we focused on whether insomnia in the elderly is more likely to cause HF, we were surprised to find that there was no statistically difference between subgroups of different age. This may due to, firstly, while total sleep time (TST), sleep efficiency, and deep sleep (slow wave sleep) decrease with aging, the elderly may accept these noticeable sleep changes as a part of normal aging owing to an adjustment of their perception of “acceptable” health with aging ^34–36^. What’s more, most age-related changes in sleep are stable after 60 years of age among older adults with excellent health ^37^. Secondly, even though more frequent arousals were found in healthy older adults than young people, older adults maintained their ability to reinitiate sleep and fell back to sleep as rapidly as younger adults ^38^. Thirdly, several studies have found that older adults nap more frequently than younger and middle-aged adults ^39^. Within the older population, studies found that nap frequency increased with age, older adults may spend less time on work and physical and social activities, and thus have more opportunities to nap than young and middle-aged adults during the day ^40^.

The results of many studies have suggested that women are more prone to insomnia than men ^16,41^. A sex difference in the association between insomnia and HF risk appears to be plausible. However, gender-based subgroup analysis of our meta-analysis showed no significantly difference between men and women. We speculate that this may be due to women have a lower HF risk at baseline compared with men. For men more often suffer from pre-existing CHD and impaired left ventricular function, whereas women more often suffer from hypertension and tend to present with sustained left ventricular function ^42^. However, given there are only two studies classified as male group in our subgroup analysis, further investigation is necessary to validate the observed sex differences in the research.

Chronic insomnia is common among patients with HF and contribute to fatigue and poor function ^43,44^. Researchers showed that cognitive behavioral therapy (CBT-I) was feasible and highly acceptable to patients with stable HF and resulted in large improvements in insomnia and fatigue ^45^. Similarly, researchers demonstrated that CBT-I and brief behavioral interventions improved sleep quality among individuals with HF^46,47^. Although further research is needed, we believe that improved assessment and treatment of insomnia may become a new prevention target HF.

### Study limitations

This study has several limitations that should be acknowledged. Firstly, the assessment of insomnia symptoms varied across studies, introducing heterogeneity in measurement tools and may make it challenging to compare and generalize the findings. However, because each study included in our meta-analysis was conducted according to standardized insomnia scales or survey questionnaires, our statistical analysis is comparable. Secondly, the presence of comorbidities such as depression, anxiety, and chronic pain alongside insomnia can confound the specific effects of insomnia on the incident of HF. Moreover, as most studies focused on adult populations and were conducted in clinical settings, potentially impacting the representation of the broader population. Furthermore, the bidirectional relationship between insomnia and HF was not extensively explored, highlighting the need for further research to understand the reciprocal interactions between these two conditions.

In conclusion, our findings suggest that DIS, a common symptom of insomnia, may serve as a valuable marker for identifying individuals at risk of HF. This highlights the critical importance of early identification and intervention for insomnia in HF patients. Additionally, recognizing the bidirectional relationship between insomnia and HF emphasizes the necessity of comprehensive management strategies that address both conditions. Healthcare professionals should consider incorporating insomnia, especially DIS assessment and interventions into the care of HF patients, aiming to optimize sleep quality and mitigate HF risk. However, further research is necessary to investigate the underlying mechanisms, establish causal relationships, and develop targeted interventions for preventing and managing insomnia in individuals at risk of HF.

## Sources of Funding

Dr Ren is supported by the National High Level Hospital Clinical Research Funding (NO.2022-NHLHCRF-LX-02-0102, NO.2022-NHLHCRF-YXHZ-01, NO.2023-NHLHCRF-YYPPLC-ZR-05), Beijing Nova Program (NO.20220484171) and Beijing Demonstration Research Ward (NO.2022-YJXBF-03-03), Dr Gao is supported by the National Natural Science Foundation of China (82300439).

## Disclosures

### Ethics approval and consent to participate

Not applicable.

### Competing interests

The authors declare no competing interests.

## Data Availability

All data in the manuscript is available

## Abbreviations

CI: confidence interval
DIS: difficulty initiating sleeping
DMS: difficulty maintaining sleeping
EMA: early-morning awakening
HF: heart failure
HR: hazard ratio
MeSH: medical subject heading
NOS: Newcastle-Ottawa Scale
NRS: non-restorative sleep
PRISMA: Preferred Reporting Items for Systematic Reviews and Meta-analyses

## Notes

### Competing Interest Statement

The authors have declared no competing interest.

### Clinical Trial

The systematic review was previously registered in PROSPERO (International Prospective Register of Systematic Reviews; Study Unique Identifier: 359352)

### Author Declarations

Ethics approval and consent to participate Not applicable.

